# Patient perspectives and hospital pay-for-performance: A qualitative study from Lebanon

**DOI:** 10.1101/2023.01.06.23284267

**Authors:** Jade Khalife, Björn Ekman, Walid Ammar, Fadi El-Jardali, Abeer Al Halabi, Elise Barakat, Maria Emmelin

**Author notes:** Corresponding author (JK).

## Abstract

**Background:** Patient perspectives have received increasing importance within health systems over the past four decades. Measures of patient experience and satisfaction are commonly used. However, these do not capture all the information available through patient engagement. An improved understanding of the various types of patient perspectives and the distinctions between them is needed. The lack of such knowledge limits the usefulness of including patient perspectives as components within pay-for-performance initiatives. This study was aimed to identify and explore patient perspectives on hospital care in Lebanon, and to describe how they relate to the national pay-for-performance initiative.

**Methods:** We conducted a qualitative study using focus group discussions with persons recently discharged after hospitalization under the coverage of the Lebanese Ministry of Public Health. This study was implemented in 2017 and involved 42 participants across eight focus groups. Qualitative content analysis was used to analyze the information provided by participants.

**Results:** Five overall themes supported by 17 categories were identified, capturing the meaning of the informants’ perspectives: health is everything; being turned into second class citizens; money and ‘wasta’ (personal connections) make all the difference; wanting to be treated with dignity and respect; and tolerating letdown, for the sake of right treatment. The most frequently prioritized statement in a ranking exercise regarding patient satisfaction was regular contact with one’s doctor.

**Conclusions:** Patient perspectives include more than what is traditionally incorporated in measures of patient satisfaction and experience. Patient valuing of health and their perceptions on each of the health system, and access and quality of care should also be taken into account. Hospital pay-for-performance initiatives can be made more responsive through a broader consideration of these perspectives. More broadly, health systems would benefit from wider engagement of patients. We propose a framework relating patient perspectives to value-based healthcare and health system performance.

## Introduction

The perspective of patients within health systems has increasingly been recognized over the past four decades. This is exemplified by several landmark reports, though the translation into action has been gradual and interspersed. Griffith’s Report in 1983 urged the inclusion of public opinion and perception of healthcare for the UK National Health Service reforms [1]. In 2001 the US Institute of Medicine (IOM) highlighted patient-centeredness as one of the six dimensions of quality of care. In 2018 the IOM also called for an expansion towards person-centeredness, whereby the care provided is “respectful of and responsive to individual preferences, needs and values” [2, 3].

At the 69^th^ World Health Assembly (2016) member states adopted the Framework on Integrated People-centered Health Services, whose vision emphasized the patients’ role in defining their needs and co-producing health services [4]. A 2018 joint report by the World Health Organization, World Bank and the Organisation for Economic Co-operation and Development (OECD) called patient-centeredness the core of quality, and called for the engagement of people and communities in service design, delivery and assessment [5]. Indeed, patient-centeredness has also been called “the doorway to all qualities”, not merely one dimension among others [6]. Moreover, the patient perspective has also been included as the personal value pillar in a re-definition of value-based healthcare [7]. Altogether, these developments represent a modern-day return to a fundamental aspect of early Hippocratic medicine: accompanying the patient and meeting their individual goals.

Understanding the patient’s perspective is an essential precursor to patient-centeredness. Improved understanding helps in ensuring that they can be “full partners in the service delivery design and governance and in improving their own health” [8]. Patient perspectives have been most commonly addressed through measures of patient satisfaction and experience. One application of these measures has been as components within pay-for-performance (P4P) initiatives, which have become widespread in healthcare over the past two decades. The earliest national initiatives were launched in the United Kingdom (UK) and the United States (US). In the UK the Quality and Outcomes Framework incentivized the measurement of patient satisfaction by practitioners [9]. In the US a national survey by the Centers for Medicare and Medicaid Services is used to capture the patients’ perspective on care received, and since 2012 the results are linked to incentive payments within the Hospital Value-Based Purchasing program [10].

The largest sub-national P4P initiative preceding those in the UK and US was that in the state of São Paulo, Brazil, which created Social Organizations in Health (OSS) in 1998 to manage hospitals [11]. The OSS model included a patient satisfaction component (complaints and completion of satisfaction surveys) for the performance component used to set global hospital budgets, alongside volume targets.

Patient experience and satisfaction are both recognized as multi-dimensional, but are ambiguous and under-theorized [12]. They also do not capture all the information available from engaging patients, such as the value patients attach to health. Therefore, despite their wide application patient perspectives are not well understood. There is a need for greater clarity on the different types of patient perspectives available, as well as on the distinction between them. The lack of such knowledge limits the usefulness of patient perspectives in pay-for-performance initiatives, potentially leading to unintended consequences [12]. More broadly, it also impedes the movement towards person-centered health systems.

In Lebanon patient satisfaction and experience measures have been included in the hospital P4P initiative since 2014, which is used for setting hospital reimbursement tiers by the Ministry of Public Health (MoPH) [13]. In 2019 these measures comprised a fifth of the total performance score set by the MoPH. The inclusion and weight given to this was intended to represent the ministry’s commitment to capturing patients’ perspectives, and incentivize hospitals to improve their performance. However, considering the aforementioned limitations of patient experience and satisfaction, it is unknown to what extent this represents patients’ true perspectives.

Studying how patients experience and perceive hospital care in Lebanon would contribute to the overall knowledge on engaging patients towards person-centered health systems, and help improve P4P design and identification of impacts.

The aim of this study was to explore how people with experience of being hospitalized perceive the healthcare, focusing on health perceptions, access to care, experiences of hospitalization and satisfaction of care.

## Methods

### Overall study design

We conducted a qualitative study using focus group discussions (FGDs) with persons who were recently discharged after a hospitalization. A qualitative approach was used, since it allows us to gather information directly from the perspective of patients, on a topic that is not well understood [14]. Qualitative methods may allow a deeper understanding of such perspectives, and would not have the limitations associated with structured questions that are characteristic of quantitative surveys [15].

FGDs were chosen, because we were interested in a wide range of views and experiences, and to encourage discussion and explanation of issues. It also limits the influence of an interviewer on a respondent’s comments, and allows for more spontaneous issues to arise [16]. FGDs rely on the interaction among group participants to encourage information generation on perceptions, attitudes and beliefs, while also allowing a facilitator to probe further when required [16]. This study is reported in accordance with the Standards for Reporting Qualitative Research (SRQR) [17].

### Study setting

This study was undertaken in Lebanon, an Eastern Mediterranean country with a population of Lebanon about 6.8 million people, including about 2 million refugees (most of whom are due to the conflict in neighboring Syria). About 52% of Lebanese citizens lack formal health insurance and are therefore under the coverage of the MoPH for hospitalization services. The MoPH contracts with public and private hospitals throughout the country to provide hospitalization services for its beneficiaries. Hospitals are reimbursed by the MoPH for 85% of the hospitalization bill, and 15% remaining as patient co-payment.

### Study population and sampling

We used a purposive sample of adult persons who had been hospitalized (and discharged) under the coverage of the Ministry of Public Health (MoPH) during the preceding 3 months. Persons covered by the MoPH are typically comprised of the poorest or most disadvantaged stratum, as opposed to those having health coverage from other payers (e.g. employees, students, police, army) [13]. The aim was to capture a maximum variation of experiences in this target group by including both men and women from different age groups and living in urban and rural settings. A database registering all hospitalizations under the MoPH coverage was used to contact potential participants via telephone. People were randomly selected from the database, which includes persons from all regions and hospitals (public and private) which had been hospitalized under the MoPH coverage (comprising about 230,000 hospitalizations per year), until we felt that we had reached an acceptably varied group of participants.

The final sample involved 42 participants (22 men, 20 women) with a median age of 49 years (range: 25-65 years). Participants resided in different regions throughout Lebanon, including urban and rural areas. All participants had had several instances of interaction with healthcare (including at least one hospitalization, by design). The median duration of discussions was 62 minutes (range: 37-82 minutes; see table 1).

**Table 1:** Focus group discussion characteristics.

| FGD # | Duration (min) | Participants, n | Sex |
| --- | --- | --- | --- |
| 1 | 65 | 7 | Men |
| 2 | 37 | 3 | Women |
| 3 | 64 | 4 | Women |
| 4 | 57 | 4 | Men |
| 5 | 58 | 4 | Men |
| 6 | 59 | 5 | Women |
| 7 | 82 | 8 | Women |
| 8 | 79 | 7 | Men |

### Discussion guide and pile-sorting statements

A discussion guide was developed using open-ended questions, to support the facilitation of the discussions. The themes of the key questions were: the meaning of health; description of local healthcare; characteristics of services received; description of ‘good’ and of ‘bad’ hospital stays; information needed upon hospital admission; and factors that would make a person revisit a hospital. The guide was piloted in one FGD and subsequently revised for greater clarity, based on the pilot results.

To increase our understanding of how patients prioritized different factors regarding their hospitalization, we also prepared a structured pile-sorting exercise. The pile sort is a method that involves asking participants to sort for example statements about the phenomenon of interest in piles of more versus less important [18]. In this study the statements were developed based on our assumptions of what factors may affect the satisfaction and experience of patients, and also using existing literature and survey tools [19, 20]. Following the end of the discussions, participants were asked to sort 16 statements into two piles (more versus less important). These included topics such as the importance of regular contact with personnel, hospital organization, cleanliness, communication, pain and privacy.

### Data collection

The FGDs were held between July and September 2017. The groups were arranged separately for men and women since we assumed that this would create a more open discussion climate.

Variability in group size was largely due to some cancellations and rescheduling of participants in the period preceding the discussions. They were all held in a private room at the MoPH headquarters in Beirut. Following an introduction to the study, all participants were asked to provide oral consent and agreed to be audio-recorded using a digital recording device.

The first author (JK) was the discussion facilitator, while two research assistants (AH, EB) noted the group layout and assessed interactions but they did not participate in the discussions. The first author (JK) introduced himself to participants as being involved in the hospital pay-for-performance initiative, which was a collaboration between the MoPH, the American University of Beirut and Lund University. The discussions involved exploring several thematic areas, with probing questions used to clarify statements or explore them in more depth. The median duration of discussions was 62 minutes, with a range between 37 to 82 minutes.

No material or financial compensation was given to participants, but all were offered complimentary transport by taxi from their residence to the discussion site and back, and snack refreshments after the FGDs had ended.

### Data analysis

The recordings were transcribed and translated from Lebanese Arabic to English verbatim (AH, EB). During the transcription process participant names and other personal identifiers (e.g. residence) were removed. Accuracy and sense-making of the transcription was iteratively assessed by the two research assistants and the facilitator.

Qualitative content analysis based on Graneheim & Lundman (2004) was used. The unit of analysis was the focus group discussion transcripts. All transcripts were read several times before coding was begun, to increase the understanding of the issues raised, as well as their depth and breadth.

Statements were maintained uncondensed, and then labelled with codes. The statements were analyzed in relation to the specific research questions that also formed the basis for the content areas. Codes sharing communalities were used to construct categories based on expressed and explicit (manifest) meaning. The latent meaning of categories was subsequently used to develop themes. An example of the analytical process is found in Table 2. NVivo 12.0 software was used to support the coding procedure.

**Table 2:** An example from the analysis process, moving from text, to code and category.

| Text | Code | Category |
| --- | --- | --- |
| You have nothing even if you don't have health, even if you own the whole world. (FGD1-P5) | Without health we have nothing | More important than money or wealth |
| Briefly, health is the whole life, who does not have good health, has nothing because the sick person is always depressed. (FGD3-P1) | Without health we have nothing |  |
| Health is everything, if you have all the money in the world but you have poor health, it means you are poor and you own nothing. (FGD2-P3) | Health is more important than money |  |
| At the end you give priority to health over other needs, this is how I think, for example I buy anything cheap, but I don't buy a cheap medicine to save money, and same for the doctor [...] a person should be frugal on everything except on his health, this is how I think [...]. (FGD3-P3) | Being frugal except with your health |  |

### Ethical considerations

The research protocol approval was granted by the Institutional Review Board (IRB) at the American University of Beirut (ID: FHS.FE.21). All participants were given oral information at the initial telephone invitation (by AH, EB) and again on-site at the introduction to the FGDs (by JK). This information included the purpose of the study; the fully voluntary nature of their participation and right of refusal; that the decision/participation would not be associated with or affect their MoPH coverage in any manner; that all material would be handled confidentially; and no results would be presented allowing participant identification. Participants were given the choice to allow the discussions to be audio-recorded or not (all accepted recording). Participants were also asked to respect that “what’s said in the group stays in the group”, since the researcher can only promise confidentiality on behalf of the research group.

## Results

The analysis resulted in five overall themes reflecting the underlying meaning of the discussions, supported by 17 categories giving the more manifest level of the interpretation. The overall results are presented in Figure 1 in relation to five content areas, based on the focus of the study. The first theme “Health is everything” indicates the high value that participants put on health. The second theme “Being turned into second class citizens” illustrates the inequity participants perceived that characterized the health system in general, while “Money and ‘wasta’ make all the difference” specifies how this inequity is manifested in the actual access to healthcare access. The fourth theme “Wanting to be treated with dignity and respect” relates directly to what participants value most when assessing hospital care while the fifth theme “Tolerating letdown, for the sake of right treatment” implies what takes precedence in the end is the medical results.

**Figure 1:**
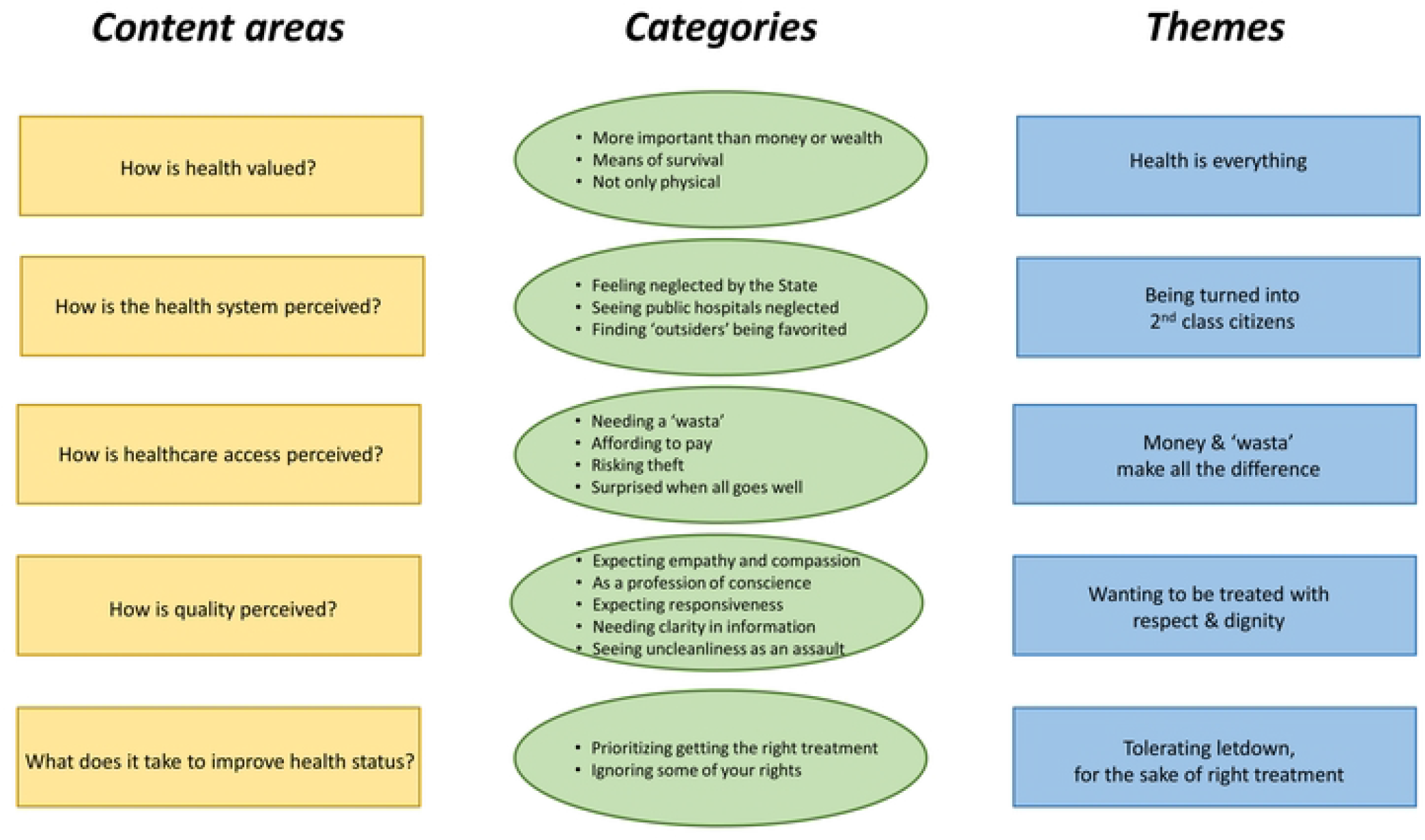
Overview of the main findings including content areas, categories and themes.

The section below presents the themes as headings in bold, while categories are in italics in the running text. Where relevant, quotes are included in italics, to support the analysis using the participants’ own words.

### Theme 1: Health is everything

This theme illustrates the valuing of health. Participants described health as everything, more important than anything else in life. Health was portrayed as **more important than money or wealth**, and they would exchange anything they own to regain their health. Health formed the crucial foundation for life. It was also considered a priority above other goods or services, whereby one may be frugal with other items, but not with health services or medications.

> *“Health is literally all life, if you don’t have health you don’t have a life; that’s it. Regardless of what your disease is, even if you only have headache, this is pain, and no one feels the pain except the patient himself*.*” (FGD1 Men-P3)*

Being healthy was not only for one’s own self, but was also important to be able to fulfill one’s role in the family and in the community. This meant supporting those dependent on you, and as a **means of survival** to be able to work and provide.

> *“Health is everything, I am a carpenter and I am paid on a daily basis, I have stopped working since a month and a half. Health is the basis of our existence, if we are not healthy we cannot work or do anything else*.*” (FGD4 Men-P3)*

Participants valued **not only physical health**, but also psychological and emotional well-being, which are important wherever a person is, be it their workplace or with their family. If you are not healthy, you also lose self-confidence. Although very important, health was also often neglected, as people get busied by other things in life and forget to take care of themselves, until they need a hospital.

> *“A person runs and endures, endures, endures and endures, and when time comes to relax, he finds himself unproductive because of his health, or could not anymore live the way he expected to live*.*” (FGD1 Men-P4)*

### Theme 2: Being turned into second-class citizens

This theme relates to how the health system is perceived. Participants described how they felt like second-class citizens in their own country. Participants recognized that **“some hospitals are not for us”**. Some attributed this to which hospitals they had access to, while others perceived they were taken advantage of by hospitals, due to their educational status. They also reported that very often the first question they would be asked at the emergency room was whether they are under the coverage of private insurance or the NSSF. Participants recognized that those with greater financial means had much better access to healthcare than the poor.

> *“The hospitals in Lebanon are classified into classes, if you tell someone you are going to [well-known hospital X], they tell you ‘this is not for you’; this is the way they reply*.*” (FGD1 Men-P1)*

Participants **felt neglected by the state**, with citizens under the coverage of other payers (non-MoPH) having better healthcare access and services. They highlighted such preferential behavior particularly being given to those covered by the NSSF and private insurance. This was perceived from the first moment of interaction between patients and hospitals; usually the hospital admission desk, but even at the emergency rooms.

> *“My parents are old and don’t have NSSF nor insurance. [My father was ill and] the hospital asked for 650 USD, and I don’t have money. I borrowed the money from someone. This is how we live in Lebanon*.*” (FGD8 Men-P6)*

A common perception was that hospitals had less respect for patients covered by the MoPH, as well as the ministry itself. They described the ministry as being weak, and they shared the excuses they would be given by hospitals for refusing hospitalizations, such as no beds being available.

Participants thought that the MoPH should more strongly advocate for and safeguard the rights of the poor and “make us feel that we are human beings”. They also wanted to see the ministry actively evaluate and regulate hospitals and primary care centers. This included investigating how some wealthy persons are reportedly receiving services under MoPH coverage, while some of the poor do not. Participants generally did not have complaints regarding MoPH requirements for hospitalization or treatment, but this was not shared by some participants who considered themselves unfairly disadvantaged. Participants recognized the adverse role of what they considered was excessive bureaucracy, such as needing to travel to different locations to receive approvals for certain procedures (e.g. to the MoPH headquarters).

> *“If you want to benefit this country you need to think of cutting down on the processes; the current way is very tiring*.*” (FGD4 Men-P4)*

Participants reported **seeing public hospitals neglected** and having limited resources devoted to them. This neglect was translated into worse services and patients being treated with less dignity and respect than those going to private hospitals. They considered that public hospitals have a major role in supporting the poor, yet their potential was ignored. This neglect was also transferred to nurses, physicians and other staff at public hospitals.

> *P4: “If they improve the services of the public hospitals, there will be no need for private ones. We have a public hospital which is the largest hospital in the Middle East […], but you cannot go there*.*”*
>
> *Moderator: “Why?”*
>
> *P1: “Because you would die!”*
>
> *P4: “First the treatment is very bad, and the cleanliness is worse!” (FGD3 Women)*

Some participants proposed solutions to improve public hospitals. Some ideas shared were: increasing funding of public hospitals, allocating the better/best doctors to public hospitals (“we always follow the big names”), improving services, and accountability of hospital directors.

Participants often avoided primary care centers. They perceived these centers to be under-staffed, and that patients had less time, and with less qualified doctors (compared to private clinics). They recognized that healthcare staff at these centers were underpaid, although they also have a duty because “there are also poor people who must be treated”.

Participants also said they **found ‘outsiders’ being favored** with faster and free treatment. This referred to refugees with coverage from international non-governmental organizations and agencies.

### Theme 3: Money and ‘wasta’ make all the difference

This theme illustrates how participants perceived access to healthcare. Participants predominantly linked this closely with the ability to have a ‘wasta’ (‘I know someone’) or have enough money.

Participants described that whatever the obstacles to access may be, one who pays directly out-of-pocket or has a **‘wasta’** would solve them. **Needing a ‘wasta’** was recognized as a major factor in accessing healthcare, across the services spectrum. This particularly included hospital admission and surgical operations, but also medications for cancer and other chronic conditions.

‘Wasta’ was also important for healthcare institutions who are connected to political or religious figures/authorities. Participants considered that this relation functioned in both directions: for healthcare institutions to be secure from accountability, and for figures/authorities to use their influence to facilitate coverage for patients at the healthcare institutions.

> *P1: “There are connections and some people may be protected by others”*
>
> *P3: “There are ‘wasayet’ all over the country”*
>
> *[…]*
>
> *P2: What is the relationship between hospitals and politics?*
>
> *P1: I will tell you: ‘hospital X’ is for ‘politician A’, hospital Y’ is for ‘political party B’, hospital Z is for ‘religious council C’” (FGD2 Women)*

Another function of ‘wasta’ was to decrease the hospital bill of patients, through the connection with political or religious figures. Participants reported several instances where their use of ‘wasta’ for themselves or relatives resulted in a considerable decrease in their hospital bill. ‘Wasta’ was also used to remedy perceived injustice or theft by the hospital through its physicians or administrative personnel. They also recognized that knowing a connection in the ministry gave them an advantage in confrontations with hospitals. Also, in some instances, the treating physician was a friend of a patient, and they would intervene with a ‘wasta’ on behalf of the patient to lower the bill.

While ‘wasta’ was considered widespread, participants recognized that this was a negative factor, although it was sometimes unavoidable to resort to. They thought that the health ministry should support them against such practices, and that getting rid of ‘wasta’ everywhere would actually improve both hospital and the health system.

> *“As long as a person wants to be admitted through ‘wasta’, the hospital will not work properly*.*” (FGD3 Women-P3)*

Not all examples of connections were perceived negatively. Living near healthcare institution sometimes gave a relation of familiarity between patients and personnel. Some participants reported having very positive interactions with personnel in local hospitals, many of whom were either relatives or neighbors, or came from the same village or town. This was also sometimes reflected in more trustful behavior regarding payment, for example by patients being allowed to ‘pay later’, as they were known to the personnel. However, there were also the fewer instances where a participant reported being taken advantage of by personnel who were relatives or neighbors.

The financial cost of health services was a major concern for participants, specifically **affording to pay**. This also affected the perception and behavior of participants towards healthcare. Hospitals were perceived by participants to pursue money above all. Patients that are able to pay out-of-pocket would receive the best treatment, and sometimes you can only be hospitalized if you have the money.

> *“We ask God to stay healthy because we do not have money to pay for healthcare services*.*” (FGD7 Women-P2)*

Money was seen as a solution to any problem encountered at hospitals, especially if one lacked a ‘wasta’. Patients reported that almost all problems occur either at admission or at the cashier. Sometimes hospitals would claim that no hospital beds are available, to deny admission for those covered by the MoPH. Some participants suggested this was a deceptive practice to allow hospitals to retain more profitable patients covered by other payers, or to compel MoPH-covered patients to pay out-of-pocket. One anticipatory approach mentioned by a participant was to claim to pay out-of-pocket at their initial interaction with hospital personnel, but after confirming bed availability he would then seek health ministry coverage.

> *“[…] I told the nurse my mother is not the daughter of a minister or a president; I cannot pay [out-of-pocket]. Then we took her to another hospital […]*.*” (FGD3 Women-P4)*

Unaffordability led some patients to early hospital discharge or to forego medical tests. Participants recognized this was harmful to their health, but they had no alternative that would allow them to pay for these services. Some participants recounted a family member pretending to be better to be discharged earlier, due to the mounting hospital bill. Others questioned the utility of doing medical diagnostic tests, reasoning that since they cannot afford treatment, then it was better for their mental health not to know more about their illness. Some participants resorted to selling personal belongings to cover costs of medical tests and treatment, and sometimes had to forego necessary medical testing for themselves or their children, for conditions such as cancer and neurological illnesses.

> *“When I had breast cancer […] I couldn’t do regular tests for checkup. I went through very hard times to do the tests and get the treatment. I sold my wedding ring [to get treatment]. The ministry couldn’t cover all the expenses; I reached a very difficult situation*.*” (FGD7 Women-P1)*

Resorting to borrowing from family and relatives was often a necessary measure, when the cost of healthcare was unaffordable for patients, despite the partial coverage by the MoPH. Sometimes this also necessitated a ‘wasta’. But recurring costs such as those for chronic medications remained a burden. Participants emphasized that they have a right to health, but they do not know how to realize these rights.

The unemployed were particularly vulnerable. Without a source of income, these persons could not afford to pay for visits to doctors’ clinics (outpatient), not afford some of their medications for chronic conditions. The safety nets available for hospitalization (e.g. MoPH coverage) were not available for patient follow-up after leaving the hospital. The impact of unaffordable healthcare costs was not limited to patients alone. Family members would be actively engaged in collecting funds to cover hospitalization costs, as well as in gathering and submitting administrative papers for coverage approvals. This sometimes meant skipping university classes or work.

**Risking theft** when seeking healthcare was emphasized by participants. This was a major concern affecting their perception of hospitals as well as health professionals. Although services were often considered to be of good quality, the lack of information and transparency over hospital bills contributed to a feeling of patient distrust towards hospitals; as though ‘they were stealing from us’. Such practice took on different forms, and could involve different actors. A common complaint from participants was of a doctor or nurse misinforming that a procedure was not covered by the MoPH. Some participants also reported doctors soliciting bribes for signing admission approval papers for patients.

> *“[…] Then we knew that the ministry does cover the surgery, although the doctor has told us that it does not […] my father stopped the cheque […] The papers were signed after ‘wasta’ […] The doctor had lied to us; why did he do that?” (FGD7 Women-P5)*

Participants noted that it was common to be asked by hospitals or physicians for payment above the MoPH pre-defined co-payment amount, sometimes by several times more. However, many were unaware that this was an illegal practice according to the contracting terms between the ministry and hospitals. In some instances, over-charging on co-payment was not hidden from patients, and hospitals or doctors attempted to justify this. Participants recognized that not all doctors are the same. Some were more helpful than others, in informing patients of their rights under MoPH coverage and encouraging them to stand up for their rights. Also, some participants were surprised by the large differences in the cost of some surgical procedures between comparable hospitals, which they considered a signal of over-charging. Other participants noted instances where their copayment was high enough to cover most or all of a surgical procedure’s cost.

Some participants reported not being informed of costly tests or procedures in a timely manner. Once hospitalized patients would find these services unaffordable. A common response of hospital personnel in such instances was that the MoPH reimbursement to hospitals was insufficient to cover hospital costs.

Insufficient information regarding payment meant that patients had a weaker role in their interaction with hospitals. Participants were aware that they were the weaker of the three parties involved (MoPH and hospitals being the other two), and that they would sometimes bear the burden of mistakes made by the hospital. In some instances, this resulted in patients being over-charged.

They were also aware of some of the limitations regarding hospital reimbursement from the MoPH. A downstream impact of these may be further over-charging on patient co-payments.

> *“Every patient admitted under the ministry’s coverage doesn’t know how much they are expected to pay […]. The ministry delays its payments to hospitals, so [hospitals] want to benefit from another source*.*” (FGD4 Men-P3)*

Some participants noted that the actual payment and invoice amounts can differ, but they would be obliged to accept it to receive treatment. In some instances, at discharge, participants would find themselves placed in an uncomfortable and embarrassing position by hospital personnel requesting payment for tests or services they claimed were not covered by the MoPH. Participants recognized the importance of speaking up about their challenges for healthcare access, especially hospitalization and medication costs. While they appreciated having coverage from the MoPH, it was far from sufficient for some.

Many participants also reported positive interactions with healthcare, whether with the ministry or hospitals. They were **surprised when all goes well**, especially when the administrative process for admission approval went smoothly; upon receiving coverage by the MoPH (85%) for their hospitalization costs; as well during hospitalization. Participants did not have to resort to neither ‘wasta’ nor over-charges for their healthcare services in these instances.

Some participants reported being denied surgery or hospitalization under the coverage of private insurance companies they were subscribed to, and then being surprised to have such services under MoPH coverage. Such interactions affected the perception and trust of participants towards the MoPH. In some instances, this was preceded by being let down due to exclusions by private insurance. Although this resulted in positive perceptions towards the health ministry, participants doubted they could rely on other ministries for delivering on other services; “no one cares about us regarding other issues”.

### Theme 4: Wanting to be treated with dignity and respect

This theme portrays how patients perceive the quality of care received at hospitals, expressed as wanting to be treated with dignity and respect, often implicitly but also explicitly. It also illustrates how patients view the health profession in itself. Reflecting on their past experiences, participants acknowledged both positive and negative interactions. These were not necessarily tied to the bio-physical outcome of treatment, but they did have an impact on how participants perceived hospitals.

**Expecting empathy and compassion** during their interactions with hospital personnel was very important. This included the manner of communication between health professionals and patients. The ease of obtaining admission approval and navigating administrative steps once hospitalized were crucial. Negative interactions had strong impressions on patients and their feelings of self-worth. They also had differing perceptions of private and public hospitals, with the former being considered to treat patients with more dignity and respect.

> *“They are putting me in an endless circle, and ultimately I am not getting anything out of it. How can I get admitted to a hospital with dignity?” (FGD1 Men-P2)*

When asked what factors result in a positive experience during a hospital stay participants often referred to past interactions when healthcare personnel had been compassionate in their behavior towards them, especially when they had been in pain. The positive demeanor and care of nurses was particularly recalled by some participants.

Participants also recounted instances when doctors decreased patient co-payments (e.g. changing hospitals, ‘wasta’), which influenced their perception of their doctor as a compassionate one. In one instance, a participant shared his story of being operated on and followed up by a physician without being asked for any payment.

> *“[The doctor said I needed surgery] and he visited me in the morning, and noon, and night. […] He doesn’t take a Lira from me. He knew I am poor and suffering. I told him I’m a farmer […] He said ‘my brother, this is helping someone in need*.*’” (FGD8 Men-P4)*

Seeing health practitioners belonging to **a profession of conscience** was also an expression of patients’ desire to be treated with dignity and respect. They considered that being humane was the most important attribute of a doctor or nurse. Participants also noted that all personnel working in healthcare institutions should be bound by the purpose of the institution, which they regarded as to help those in need. This was also specified for administrative personnel, including those in hospital admission and cashier roles.

> *“Humanity is the most important thing to be found at hospitals” (FGD2 Women-P1)*

This also meant that the over-arching priority should be ensuring every person’s ability to receive care, regardless of the ability to pay. This was considered as a fundamental right for humans, despite many being deprived of it.

Participants were also **expecting responsiveness** from staff while hospitalized, and reported both positive and negative experiences. There was understanding regarding the long working hours and challenging conditions that hospital staff worked under, particularly for nurses. Some suggested this to be a reason why responsiveness was sometimes lacking. More ‘difficult’ or demanding patients were also considered to be a challenge for staff. However, maintaining patient-centeredness was considered a necessity.

*“When the nurse is in this profession, he must be expecting what he will face, he must not get annoyed and he must be patient. When the patient is at the hospital, he isn’t going to be faking it, he will really be in pain. This is why he will be nagging; because no one nags for no reason*.*” (FGD7 Women-P5)*

Staff responsiveness was particularly important when a patient was in pain, or needed aid to use the toilet. The lack of responsiveness in such situations led to strongly negative experiences by patients.

Participants highly valued the time personnel devoted to them. It was especially important to have enough time with the doctor, in order to have a clear explanation of their medical condition and treatment options, as well as to get responses to their questions. Doctors that did not make time for their patients were perceived as unresponsive and arrogant, regardless of their reasons.

> *Moderator: “How would you differentiate between a good and humane doctor, and a bad one?”*
>
> *P4: “When he provides you with information, as I told you. Because my doctor’s clinic is so busy, if I want to ask him a question he says ‘there is no need to know about these things, I know about them’; this annoyed me*.*” […]*
>
> *P5: “The doctor is good when he gives you from his time, even though sometimes he is in a rush, but he has to make you relaxed, to explain your condition to you*.*” (FGD7 Women)*

It was also important to have a doctor you are comfortable with. Participants highlighted the importance of having a doctor they could rely on. ‘Following the doctor’ was how participants largely explained their decision to visit or re-visit a hospital. This sometimes included situations where they were not comfortable with the hospital. Participants were aware that countries would have a mix of better and worse doctors, and that none could be right or perfect all the time. They also acknowledged that chance also plays a role in whether you find a good doctor or not, as well as the importance of hearing the recommendation of friends or relatives before choosing a doctor.

**Needing clarity in the information** provided by both hospitals and the MoPH was important to participants. From hospitals, they expected more clarity on the treatment options and how long their stay may be. They highlighted the need clearer information from both the MoPH and hospitals regarding the amount for co-payment. More broadly, participants wanted to be more aware of their rights through the MoPH, and thus more empowered to defend themselves.

> *“The ministry must improve citizens’ awareness […] We should know which hospitals we cannot be admitted to, the services and benefits we can get […] When we are aware, we can fight for our rights*.*” (FGD 7 Women-P5)*

Participants emphasized that information clarity is perhaps even more important in health settings than in other (non-health) services since as a patient one is more vulnerable and dependent on others.

More broadly, participants thought it was important to know which hospitals were better performers; the location and medications accessible from medication dispensaries and primary care centers; and the cost (or co-payment) of surgical procedures under MoPH coverage. Regarding the latter, it participants found it illogical and a lapse of accountability that co-payments could not be pre-determined and committed to. Many were unaware of the difference between a deposit receipt and their hospitalization bill, as well as their right to have a detailed hospital bill. There was minimal information provided at discharge regarding their bill, most of which was verbal, not written, with the exception of the receipt.

Cleanliness was an important issue for participants, particularly regarding toilets and bedsheets. For some, this was more important than the medical treatment. Participants described some of their experiences and how they **saw uncleanliness as an assault** and as a danger to themselves.

> *“I was worried of getting a virus there; the toilets are not clean, such things makes me worried of getting an infection […] hygiene and toilets are very important. I care about such things more than treatment […] cleanliness is the most important factor*.*” (FGD7 Women-P5)*

Cleanliness was also something visible that patients sometimes directly associated with quality of treatment. Participants noted cues they would use to assess cleanliness. These included spotting spider webs on ceilings, and the frequency and timing of cleaning staff work. Cleanliness left a strong impression of hospitals among participants; “you see and sense cleanliness”.

The responsiveness of nurses and cleaning staff after an incident was also important. Participants with a positive experiences appreciated being attended to quickly, while others recalled negative experiences after having to wait a few hours for a change of bedsheets soiled by their surgical drainage.

> *“They treated me well […] my legs were swollen and I couldn’t step on the floor; I couldn’t use the toilet. They used to be next to me after one minute of ringing the bell to clean me up and wash me*.*” (FGD5 Men-P3)*

### Theme 5: Tolerating letdown, for the sake of right treatment

This theme illustrates what participants see it takes to improve one’s health status. They underscored that their purpose in being in a hospital was **to get the right treatment**, and that they generally prioritized this above all else. This was also a major reason why a patient would consider re-visiting a hospital. Some would accept being in a less-favored hospital, if it meant they could ‘follow their doctor’ and get appropriately treated.

> *“If nurses have a disagreement, it would affect you […] Sometimes nurses are very nice, it depends on your chance […] At the end we say it is fine and we thank God there is a hospital that admits us*.*” (FGD4 Men-P4)*

Some participants noted that they tolerated some delays or behaviors, because they empathized with healthcare personnel’s working conditions. Participants also suggested personnel should have shorter working schedules and more rest time.

Getting the right treatment sometimes involved **ignoring some of your rights**. Participants would sometimes be compelled to not voice their concerns or displeasure towards personnel, because they did not want to compromise on their treatment outcomes. In some instances, participants would try to overlook negative behaviors or incidents, and focus on having their health status improved.

> *“I ignore lots of things, you can say that I ignore 40-50% of my rights, the most important thing is to get the treatment*.*” (FGD4 Men-P4)*

### Pile sorting exercise

When asked directly to rank the 16 pre-formulated statements on patient satisfaction, the most frequently prioritized statement was regular contact with one’s doctor, with nearly all participants considering this to be more important. This was followed by clear medication and care instructions at discharge, room cleanliness, shared decision-making, and good hospital organization (see table 3). The least frequently prioritized statements were regarding food, privacy and ability to discuss fears and anxieties. However, it was notable that even these statements were still considered ‘more important’ by about half of the participants.

**Table 3:**
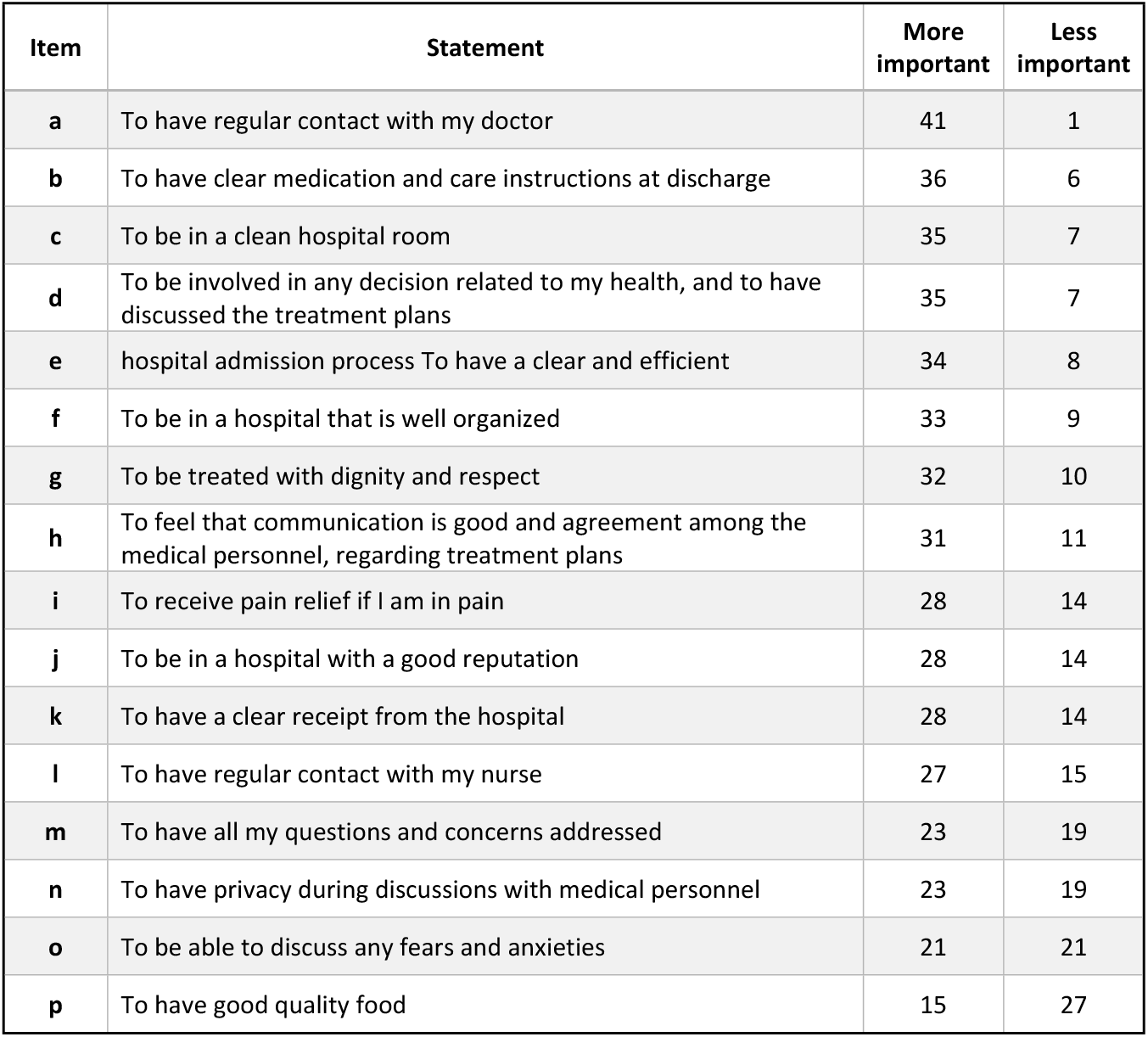
Results from the pile sorting (n=42)

Statements were similarly prioritized by women and men participants, with few exceptions. Women more frequently considered it more important to be able to discuss any fears and anxieties (13:8), and to have a clear receipt from the hospital (16:12). Men more frequently considered it more important to have regular contact with their nurse (16:11), and to receive pain relief if in need (16:12).

The results of the sorting exercise reflected the wide range of factors that patients consider important during their hospitalization and affects their satisfaction. This provided ‘patient satisfaction’ as a sixth patient perspective, following the previously identified five perspectives.

## Discussion

In this qualitative study, we explored patient perspectives on hospital care in Lebanon. All participants had been recently discharged from a hospital under the coverage of the Ministry of Public Health, which typically includes the poorest or most disadvantaged stratum of Lebanese. To our knowledge, this was the first qualitative research on hospital patient perspectives in Lebanon. It also adds to the very limited international evidence base on patient perspectives, and how patients can evaluate their care [21]. A key message of this research was that patients have a clear idea of what is needed to improve their healthcare access and experience.

### Summary of findings

Health was above everything and necessary for living and having a disease need not be accompanied by dis-ease, if well managed. The participants held the health profession as one of conscience, where all its practitioners are held to a standard. Despite the prioritization of health, they saw that being under the coverage of MoPH meant that they were turned into second-class citizens and felt neglected by the State. They wanted to see more invested in public hospitals, which they considered have been neglected, despite being a national cornerstone for supporting the ill. The results also indicate that money and ‘wasta’ can overcome any barriers to healthcare, and that this group of patients were vulnerable to over-charging of co-payments. How to pay for one’s healthcare was a major concern, causing worry for financial debt.

Participants also described positive experiences with healthcare, which were accompanied by feelings of surprise and even pride. This may reflect the wider resonance of negative experiences and barriers in shaping public perception. Health practitioners and institutions are not homogenous, and a patient may have the ‘good fortune’ to interact with those that are both professional and humane. During hospitalization patients wanted to be treated with dignity and respect. They expected personnel to be empathetic, compassionate and responsive to their needs. Cleanliness was important in affecting patients’ perception of hospitals. They wanted to have sufficient time with their doctors to better understand their health status and treatment. Patients wanted to know their rights to be able to break down barriers to healthcare, particularly knowing which services they were entitled to. They also wanted clearer information regarding their hospitalization bill and co-payment. Patients also thought it was important to know which are the better performing hospitals. Patients were sometimes let down, but they tolerated much of this, including compromising on their rights. They did this for the sake of getting treatment to improve their health.

Patients placed a distinctly high importance on the contact with their doctor, as well as hospital cleanliness. A wide range of factors were important for patients’ hospital experience and satisfaction. Some were considered more important than others, particularly having clear information at discharge, room cleanliness and shared decision-making. Also, patients’ satisfaction was not only determined by their interactions and surroundings, but also by their worries regarding payment.

### How this relates to other studies

Unsurprisingly, many of the findings were in agreement with previous studies investigating what patients perceived to matter to them. Patients want to be treated by health professionals who are humane, informative, available and not money-driven [22-26].

The purpose of being hospitalized is to receive appropriate medical treatment or diagnosis. Nevertheless, our findings suggest that other factors may be equally or more important to some patients, such as having humane personnel and hospital cleanliness. This was not unusual, considering reports such a 2018 England survey, which found that twice as many people would prioritize compassion over medical outcome, than those who would not [27]. While not detracting from the primacy of the medical outcome, this underlines the importance of compassion.

The importance of responsiveness was not limited to patient perceptions alone. It has also been linked to incidence of hospital-acquired infections, with poor responsiveness possibly acting as a symptom of wider hospital problems [28]. Participants referred to particular positive or negative experiences repeatedly, often to respond to very different issues. This suggests that discrete experiences can have a major role in shaping patient perceptions of both the hospital and the overall health system. This was also in agreement with research supporting the prominent impact of patient perceptions of care [29, 30].

Our study contributes towards untangling the patient experience from satisfaction. ‘Patient satisfaction’ has been the predominant term used to encompass the patient’s perspective since the 1970s, originating from consumerist theories as an analogue to ‘customer satisfaction’. In a healthcare context, however, satisfaction has been challenging to define. Our findings suggest that while satisfaction is important, it is a downstream result of other factors such as being comfortable with all or most aspects of care. Fundamentally, satisfaction is an emotion that refers to how patients feel. It is therefore not entirely explainable through objective reasoning. Most theories that attempted to explain satisfaction revolved around the relation with expectation [31]. Nevertheless, it is important to note that expectancy theories were insufficient, and expectation has been found to explain only a minor share of the variation in satisfaction reports [31]. Therefore, satisfaction should not be used alone, as a solitary reductionist measure, while ignoring other patient perspectives.

Patient experience is a term often used interchangeably with patient satisfaction. However, despite some overlap, the two terms are not the same. Patient experience may be defined as the sum of all interactions that influence perceptions of patients. Our findings concurred with other research in that this usually reflects the perception of the quality of care experienced by the patient [32]. Our findings also revealed that the patient experience also had a wider influence, such on perceptions regarding healthcare access or overall health system. What precisely is included in perception of quality is less clear, but can be ascertained through the questions included in different measurement tools.

### How this is useful for P4P in Lebanon and elsewhere

There are numerous tools designed to quantify the patient’s perception of quality in hospitals, some of which have been used in different pay-for-performance initiatives. A few are considered of high-quality, such as the Ethiopian Patient Experiences with Inpatient Care (I-PAHC), Indian Patient Perceptions of Quality (PPQ), and the US Hospital Consumer Assessment of Healthcare Providers and Systems (HCAHPS) [33]. Such tools are predominantly developed using expert opinion, rather than being informed directly by patient perspectives. As such, to improve the validity of these tools, it is important to use qualitative research conducted with patients themselves [26]. Quantitative and qualitative investigations are complementary. Although qualitative investigations make comparisons and generalizations difficult, their strength lies in being based on the patients’ own words [15]. Our research contributes information that may be used in improving the validity of quantitative tools used to evaluate patient perspectives.

In the Lebanese context, the tool currently used by the MoPH for its P4P patient-related component was a locally adapted and abbreviated version of the US HCAHPS. By design, this focused predominantly on perception of quality. Our research identified issues that are partly or entirely uncaptured by this tool. Some of these related to perception of quality (e.g. information clarity), but also to other patient perspectives (e.g. risk of theft). It may therefore be useful for the MoPH to consider revision of this tool to enhance its validity. We also note that a preliminary analysis of this data was used to modify some items of the MoPH tool for the 2018 patient survey (e.g. time spent with personnel, discharge information). Our findings also suggest several practical actions that can be undertaken to improve patient healthcare access and experience, such as those relating to co-payment and over-charging.

More widely, our findings may also be useful towards developing a Lebanese patient-centered health system. The high value patients attach to health is not necessarily reflected in national governance and spending. Though patients shared numerous positive experiences, our findings suggest there is much space for development of the health system, particularly towards supporting public hospitals and increased accountability of health institutions and personnel. Healthcare cannot always deliver on cure, but it should also address wider patient perspectives and not only health status. This is perhaps best expressed in the oft-cited aphorism adopted by the physician Edward Livingston Trudeau for the Saranac Lake sanatorium: “to cure sometimes, to relieve often, to comfort always”.

### How this relates to value-based care and health systems

We also sought to examine how patient perspectives relate to value-based care and person-centered health systems. We had identified six distinct but related patient perspectives. While many health systems include some measures of perception of quality, others are largely uncaptured, namely the valuing of health, perception of access and perception of overall health system. Some systems additionally measure health status, including functionality, often using patient reported outcome measures. We note that these are absent at most Lebanese hospitals.

Given the absence of a framework to relate the six patient perspectives to health system performance and value-based healthcare, we developed the framework shown in figure 2. This uses the value pillars recently proposed by the WHO EU Health Observatory and the European Commission, together with the Kruk and Freedman framework for health systems performance [7, 34, 35]. The content areas in figure 1 link to the patient perspectives in figure 2.

**Figure 2:**
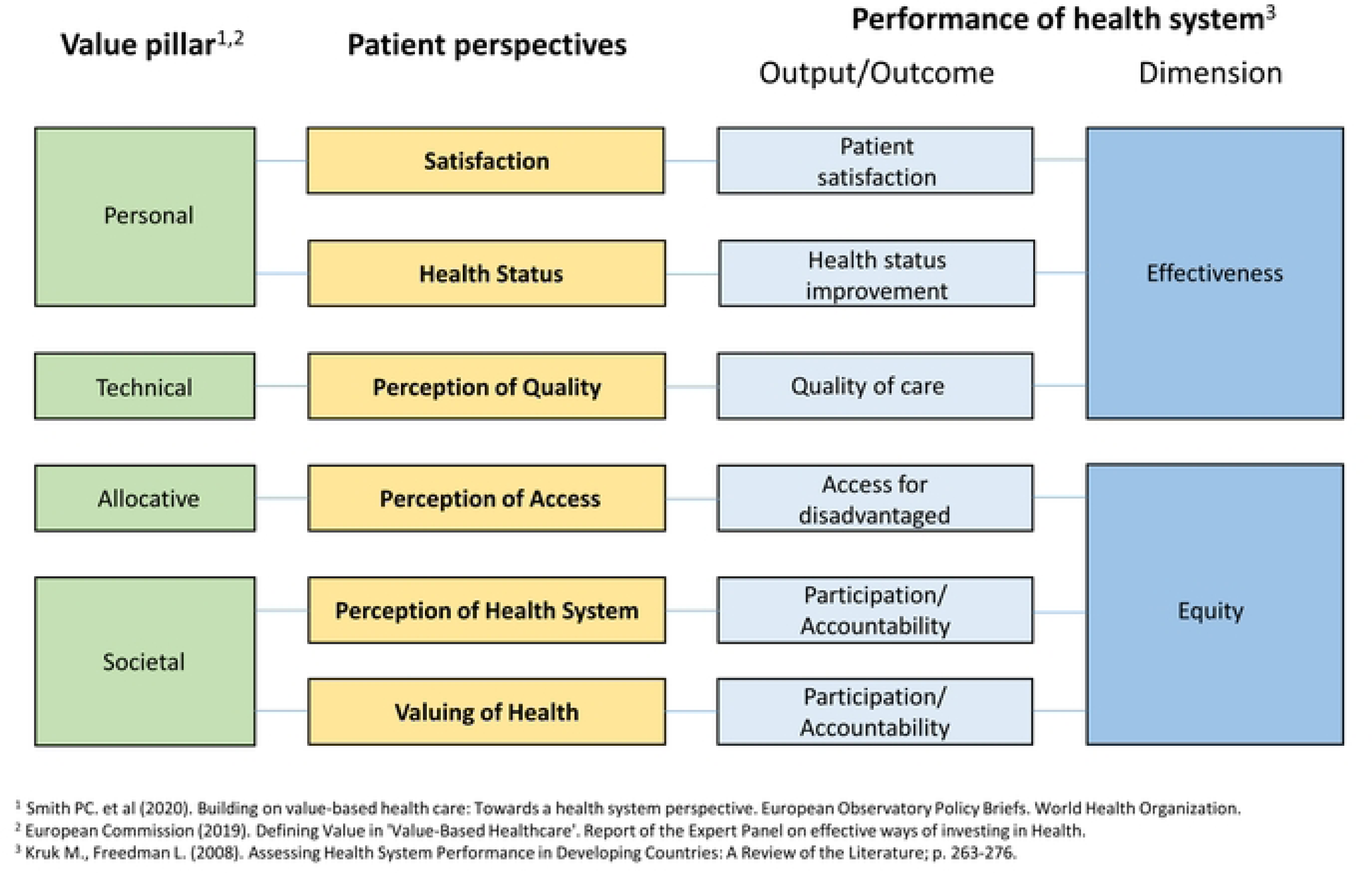
Relating patient perspectives to value-based care and health systems performance.

Besides demonstrating the non-interchangeability of patient satisfaction and experience, we also suggest this framework to clarify the current and potential role that patients can have within health systems. Engaging with patients need not be limited to measurement of quality (indirectly), satisfaction and health status. It may also involve assessing healthcare access and the overall health system, thus contributing towards decreasing inequity within populations. How patients value health is important to capture, as it relates to both accountability and participation within a health system.

Value-based care has traditionally been focused on technical value (i.e. cost-effectiveness). More recent initiatives have in addition proposed allocative, societal and personal values [7, 35]. These generally concern the equitable resource distribution within a population (allocative value); the contribution of healthcare towards solidarity, connectedness and social cohesion (societal value); and patient-centeredness (personal value). Relating these values to different patient perspectives allows value-based programs (including pay-for-performance) to more widely incentivize values including allocative and societal values which are largely unaddressed.

### Trustworthiness and methodological considerations

The ‘trustworthiness’ of our research was based on careful consideration of data credibility, dependability and transferability [36].

Credibility was improved by selecting a maximum variation of persons who had been recently discharged followed hospitalization under MoPH coverage. This sample purposefully included men and women, from different age groups and representing both rural and urban areas. Credibility was further enhanced by the first author (and moderator) having had experience with the MoPH in patient satisfaction and its P4P initiative. In addition, the moderator and research assistants (AH, EB) held debriefing sessions following each FGD [14]. The MoPH was not involved in the analysis of the data developed.

Dependability was improved by using a discussion guide, which ensured core questions were addressed, while allowing discussions to differently explore further topics.

To facilitate transferability, we have described the context of the study, the selection of participants and their general background, as well as the data collection and analytical process. We also made wide use of quotations where deemed appropriate. It is thus up to the readers to assess the relevance of the results in other settings.

Participants shared a wide range of opinions, many of which was critical, as well as very personal stories. This suggests that they felt comfortable to speak freely. We note that the discussions were held at the MoPH headquarters, and while some participants did share critical opinions also of the ministry, we cannot rule out potential impact of the host location.

## Conclusion

Patient perspectives include more than patient experience and satisfaction. In addition to traditional measures, patients may also be engaged on their valuing of health, and perceptions on healthcare access and quality of care.

The drive towards patient or people-centered health systems should incorporate a wider consideration of patient perspectives. Pay-for-performance initiatives can also be more responsive and better align patient and provider interests by a broader consideration of patient perspectives. We also propose a framework for relating patient perspectives to value-based healthcare and health system performance.

This study specifically highlights the importance of health to people in Lebanon, and the need to prioritize health services to match with people’s expectations. Patients want to be treated with dignity and respect by hospitals throughout their healthcare journey. Addressing inequity should include curbing the influence of ‘wasta’ and greater protection against financial exploitation by providers. The standardization of coverage among healthcare payers (or their unification) would circumvent patients being turned into “second-class citizens”.

Hippocratic medicine was centered on accompanying the patient and meeting their individual goals. Health systems aiming to return to this fundamental aspect should more widely engage patients for their perspectives, and incorporate these within health system design.

## Data Availability

Data cannot be shared publicly, to ensure participant confidentiality. Data are available from the Institutional Review Board at the American University of Beirut (contact ID: FHS.FE.21) for researchers who meet the criteria for access to confidential data.

## Acknowledgements

We are very grateful to all the participants in the focus group discussions, without whom this research would not have been possible. We are also grateful to following individuals at the Lebanese MoPH: Rita Freiha, Hilda Harb and Jihad Makouk (feedback and insight); and Rabiaa Rachid and Jenny Romanos (data extraction for sampling); and to Anne Mills (London School of Hygiene and Tropical Medicine, project senior advisor).

## Supplementary figures

**Supplementary figure 1:**
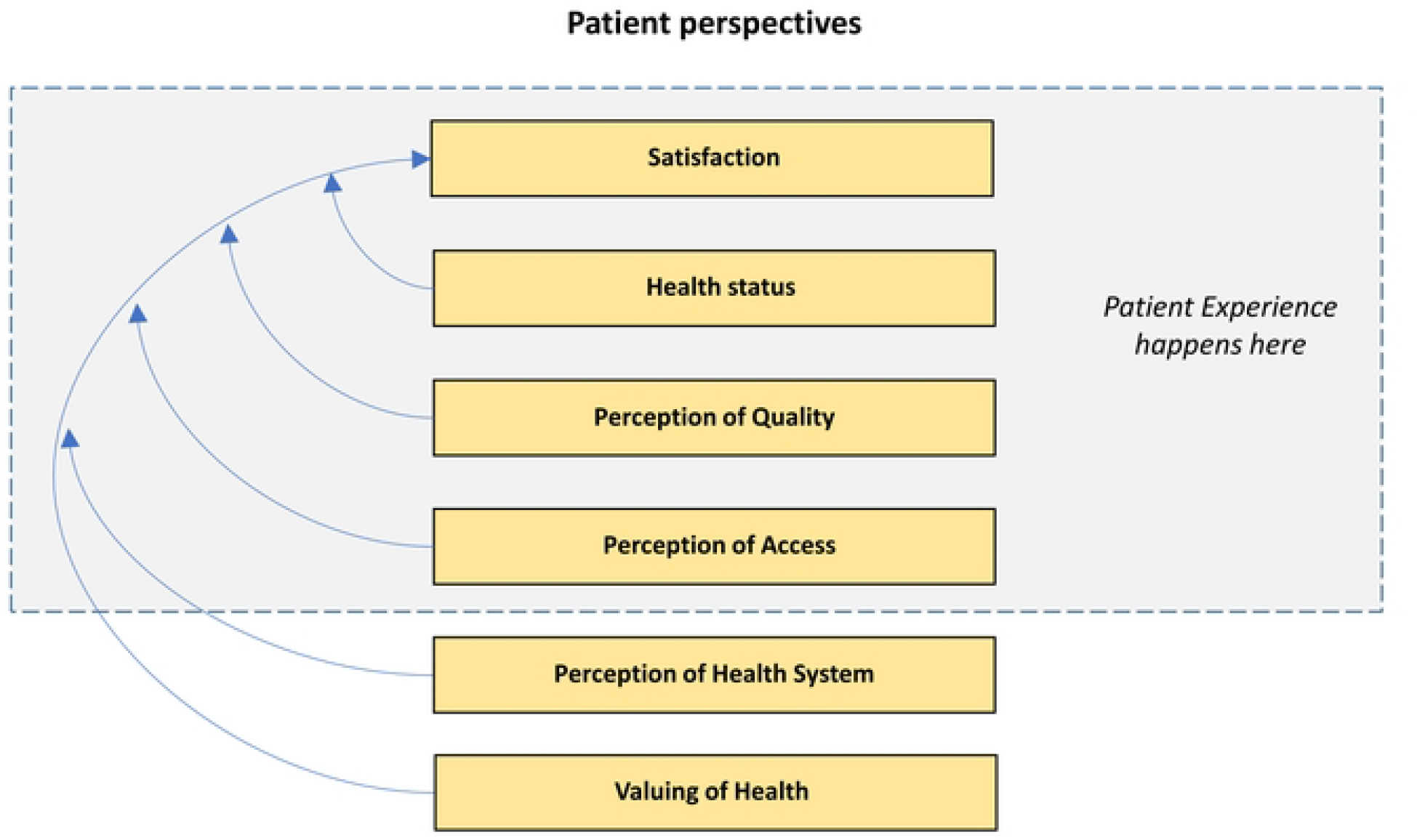
Demarcating where patient experience occurs in relation to the six patient perspectives.

### Appendix

The key questions included in the discussion guide are listed below.

Q1. What does health mean to you?

Q2. Please describe the health care offered in this area? Probe on hospitals and primary care

Q3. What characterizes the services that you have experienced so far? Probe on hospitals and primary care

Q4. How would you describe a ‘good hospital stay’? Probe on the role of different components (treatment, personal interaction, hygiene etc.)

Q5. How would you describe a ‘bad stay in hospital’? Probe the role of different components (treatment, personal interaction, hygiene etc.)

Q6. What information about a hospital stay do people need when they are admitted?

Q7. What would make a person visit the same hospital again?

